# Covid-19 is a leading cause of death in children and young people ages 0-19 years in the United States

**DOI:** 10.1101/2022.05.23.22275458

**Authors:** Seth Flaxman, Charles Whittaker, Elizaveta Semenova, Theo Rashid, Robbie M Parks, Alexandra Blenkinsop, H Juliette T Unwin, Swapnil Mishra, Samir Bhatt, Deepti Gurdasani, Oliver Ratmann

**Affiliations:** Department of Computer Science, University of Oxford, Oxford, UK; MRC Centre for Global Infectious Disease Analysis, Jameel Institute for Disease and Emergency Analytics, Imperial College London, UK; Department of Epidemiology and Biostatistics, School of Public Health, Imperial College London, UK; Department of Environmental Health Sciences, Mailman School of Public Health, Columbia University, New York, New York, USA; Department of Mathematics, Imperial College London, UK; Department of Public Health, University of Copenhagen, Copenhagen, Denmark; Queen Mary University of London

## Abstract

Covid-19 has caused more than 1 million deaths in the US, including at least 1,204 deaths among children and young people (CYP) aged 0-19 years, with 796 occurring in the one year period April 1, 2021 - March 31, 2022. Deaths among US CYP are rare in general, and so we argue here that the mortality burden of Covid-19 in CYP is best understood in the context of all other causes of CYP death. Using publicly available data from CDC WONDER on NCHS’s 113 Selected Causes of Death, and comparing to mortality in 2019, the immediate pre-pandemic period, we find that Covid-19 mortality is among the 10 leading causes of death in CYP aged 0-19 years in the US, ranking 8th among all causes of deaths, 5th in disease-related causes of deaths (excluding accidents, assault and suicide), and 1st in deaths caused by infectious or respiratory diseases. Covid-19 deaths constitute 2.3% of the 10 leading causes of death in this age group. Covid-19 caused substantially more deaths in CYP than major vaccine-preventable diseases did historically in the period before vaccines became available. Various factors including underreporting and Covid-19’s role as a contributing cause of death from other diseases mean that our estimates may understate the true mortality burden of Covid-19. Our findings underscore the public health relevance of Covid-19 to CYP. In the likely future context of sustained SARS-CoV-2 circulation, pharmaceutical and non-pharmaceutical interventions will continue to play an important role in limiting transmission of the virus in CYP and mitigating severe disease.

## Introduction

In the 12-month period April 1, 2021 - March 31, 2022, there were more than 381,000 deaths from Covid-19 in the US^1^ (a rate of 115 / 100,000 population). In children and young people (CYP) aged 0-19 years there were 796 deaths from Covid-19 reported in this time period (1.0 / 100,000 population). The overall risk of death from Covid-19 in CYP is thus substantially less than in other age groups in the US. However, deaths in US CYP from all causes are rare (49.4 per 100,000 in 2019 for 0-19 year olds; 25.0 per 100,000 for 1-19 year olds); the mortality burden of Covid-19 is therefore best understood by comparing it to other significant causes of CYP mortality from a recent pre-Covid-19 period. For this purpose, we use the US Centers for Disease Control and Prevention (CDC) WONDER database of mortality statistics on underlying cause of death. Rankable causes of death are defined by the US National Center for Health Statistics (NCHS)’s grouping of the 113 Selected Causes of Death mortality tabulation list of ICD-10 codes for underlying cause of death. These were originally drawn up in 1951 to allow comparisons for public health purposes, and are regularly updated^2^: Covid-19 (ICD code U07.1) was added to this list in October 2020^3^. We consider the 10 leading causes of death in 2019, that is, the ordered list (1st through 10th) of causes that occur most frequently among NCHS’s rankable causes of death^2^. Leading causes of death are one among various ways of understanding mortality and burden of disease. They are a starting point for a high-level understanding of public health priorities and resource allocation.

## Methods

We obtained the 10 leading causes of death among the rankable groupings of underlying causes of death from the NCHS 113 Selected Causes of Death by age group in 2019 from CDC WONDER^4^, comprehensively described elsewhere^2^. We compared these to Covid-19 mortality as an underlying cause of death in our study period, April 1, 2021-March 31, 2022, obtained from CDC WONDER Provisional Mortality Statistics^1^ (the most recent 12-month period in which data is close to complete^5^.) An underlying cause of death is defined as a disease or injury that initiates a series of events leading directly to death. To determine pediatric death rates by cause of death, we used 2021 population size estimates by single year of age from the US Census Bureau^6^. We report crude death rates per 100,000 population.

We note that NCHS’s list of rankable causes of death usually group together many individual ICD codes^2^, but Covid-19 is considered as a cause of death for a single ICD code, U07.1. Thus, we are comparing the underlying cause of death from a *single* pathogen (SARS-CoV-2) to groupings of multiple underlying causes of death (such as influenza and pneumonia). We also note that the CDC WONDER Provisional Mortality Statistics could still be revised in the future. In addition, they do not include deaths where Covid-19 was considered to have been a contributing cause of death,^1^ and thus differ from other datasets reporting Covid-19 deaths, e.g. NCHS has a separate source reporting counts of deaths involving Covid-19^7^ (a previous version of this study incorrectly used these data^8^).

## Results

Pediatric Covid-19 death rates follow a characteristic U-shaped curve across age groups in the US (Figure 1a), a commonly observed pattern^9,10^. We consider five age brackets: <1 year, 1-4 years, 5-9 years, 10-14 years, and 15-19 years. In the study period, Covid-19 death rates in infants aged <1 year were 3.9 deaths per 100,000 population. The Covid-19 death rate was below 0.5 per 100,000 in children aged 1-4, 5-9, and 10-14 years old, increasing to 1.8 per 100,000 in 15-19-year olds.

**Figure 1.**
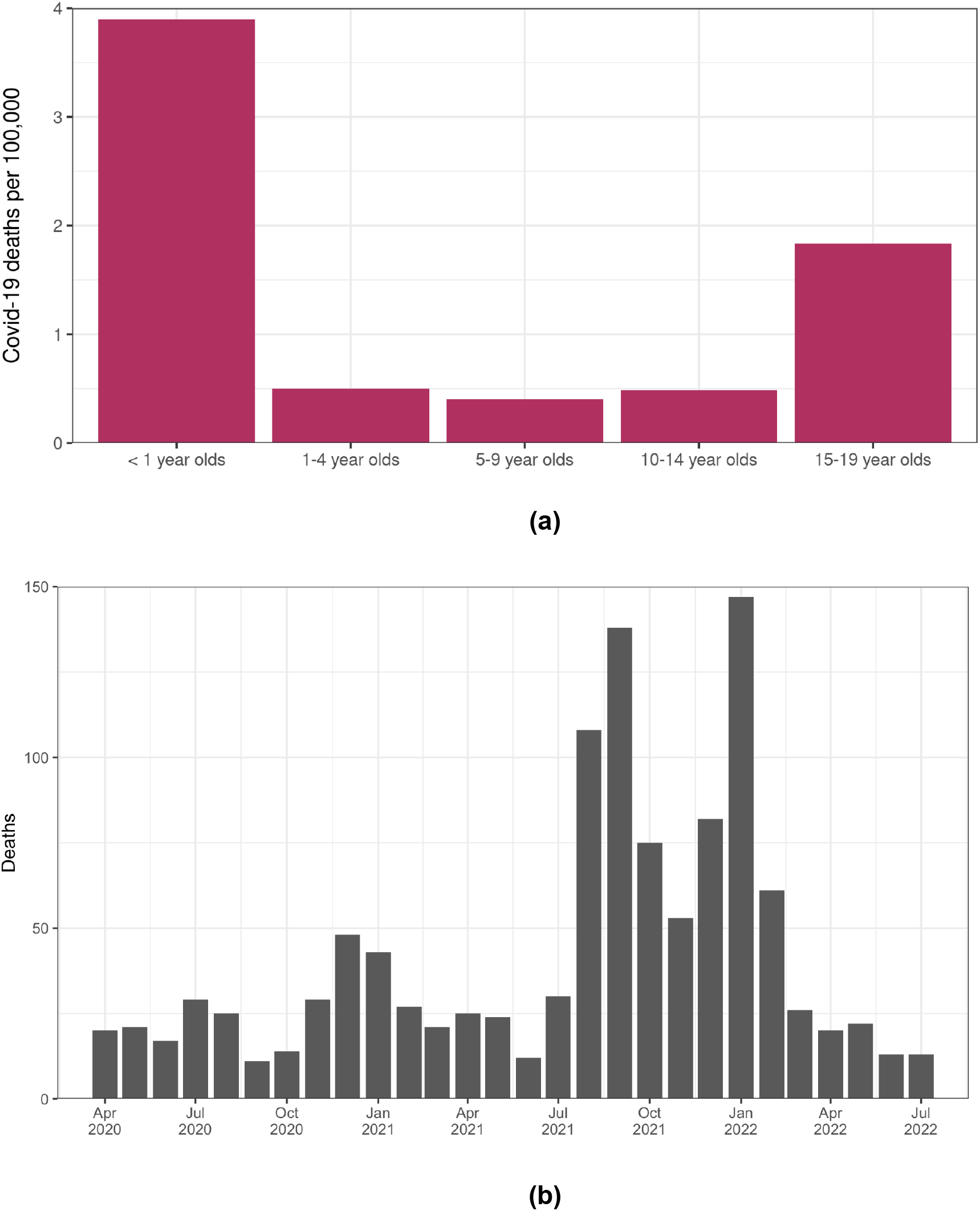
**(a)** Covid-19 death rates in the US for children and young people, where Covid-19 is listed as the underlying cause of death (ICD code U07.1) on the death certificate^1^. Rates are calculated as Covid-19 deaths for the period April 1, 2021-March 31, 2022 per 100,000 population (2021 population estimates). **(b)** Monthly Covid-19 deaths in the US of children and young people, where Covid-19 is listed as the underlying cause of death (ICD code U07.1) on the death certificate^1^.

**Figure 2.**
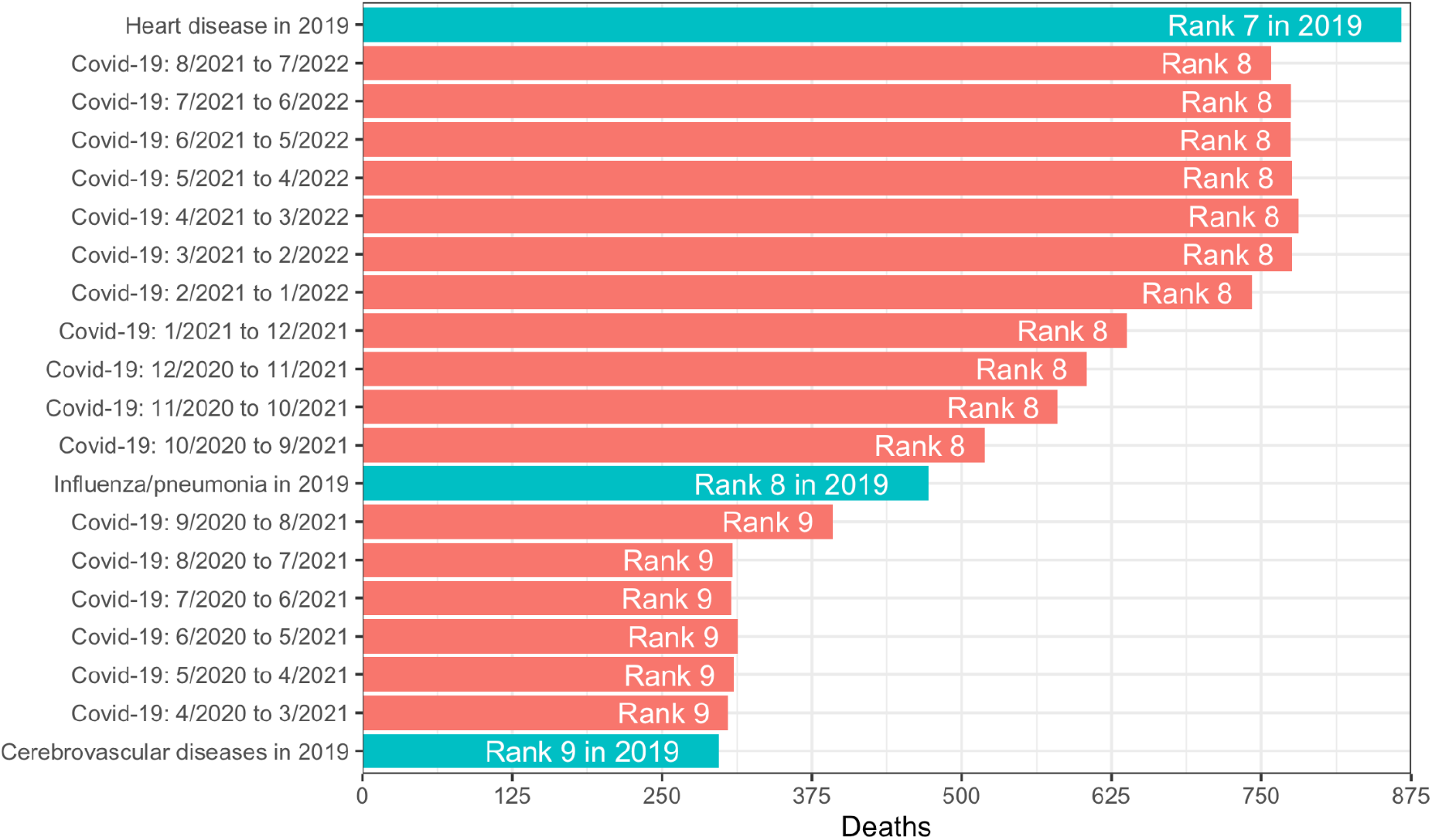
For children and young people aged 0-19 years in 2019, leading causes of death included heart disease (ranked 7th), influenza/pneumonia (8th), and cerebrovascular diseases (9th). We compare these causes of death to Covid-19 deaths in each 12 month period for which data is available: April 2020-March 2021, May 2020-April 2021, …, August 2021-July 2022.

In 2019, leading causes of CYP deaths aged 0-19 years (Table 1) were perinatal conditions (12.7 / 100,000), accidents (9.1 / 100,000), congenital malformations or deformations (6.5 / 100,000), assault (3.4 / 100,000), suicide (3.4 / 100,000), malignant neoplasms (2.1 / 100,000), diseases of the heart (1.1 / 100,000), and influenza and pneumonia (0.6 / 100,000).

**Table 1.** Leading causes of death from the rankable causes on the NCHS 113 Selected Causes of Death List, for children and young people aged 0-19 years in 2019 in the US ranked, compared to Covid-19 deaths (April 1, 2021-March 31, 2022). Deaths, crude rates per 100,000, ranks, and percentage of the 10 leading causes of death are shown. Covid-19 is the 8th leading cause of death, and the 5th leading cause of death in disease-related causes of deaths (excluding accidents, assault and suicide),

| **Table 1**<br>**Ages: 0-19 year olds** |  |  |  |  |
| --- | --- | --- | --- | --- |
| **Leading Causes of Death** | **Crude Rate**<br>**(per 100,000)** | **Deaths** | **Rank** | **% of**<br>**top 10** |
| #Certain conditions originating in the perinatal period (P00-P96) | 12.7 | 10387 | 1 | 31.7 |
| #Accidents (unintentional injuries) (V01-X59,Y85-Y86) | 9.1 | 7444 | 2 | 22.7 |
| #Congenital malformations, deformations and chromosomal abnormalities (Q00-Q99) | 6.5 | 5286 | 3 | 16.1 |
| #Assault (homicide) (*U01-*U02,X85-Y09,Y87.1) | 3.4 | 2770 | 4 | 8.5 |
| #Intentional self-harm (suicide) (*U03,X60-X84,Y87.0) | 3.4 | 2756 | 4 | 8.4 |
| #Malignant neoplasms (C00-C97) | 2.1 | 1704 | 6 | 5.2 |
| #Diseases of heart (I00-I09,I11,I13,I20-I51) | 1.1 | 867 | 7 | 2.6 |
| #COVID-19 (U07.1) | 1.0 | 796 | 8 | 2.4 |
| #Influenza and pneumonia (J09-J18) | 0.6 | 472 | 9 | 1.4 |
| #Cerebrovascular diseases (I60-I69) | 0.4 | 297 | 10 | 0.9 |

For comparison, in the study period, April 1, 2021-March 31, 2022, there were 796 CYP deaths aged 0-19 years reported whose underlying cause was Covid-19 (1.0 / 100,000), meaning it ranks as the 8th leading cause of death (Table 1) and accounts for 2.3% of the 10 leading causes of death. Rankings disaggregated by age group are shown in Table 2, demonstrating that Covid-19 ranks consistently in the 10 leading causes of death in CYP in all age groups. Ranks are: 7th (<1 year olds), 7th (1-4 year olds), 6th (5-9 year olds), 6th (10-14 year olds), and 5th (15-19 year olds). Covid-19 accounted for 0.8% (<1 year old), 2.8% (1-4 year olds), 4.5% (5-9 year olds), 4.1% (10-14 year olds), and 4.2% (15-19 year olds) of the 10 leading causes of death by age group.

**Table 2.**
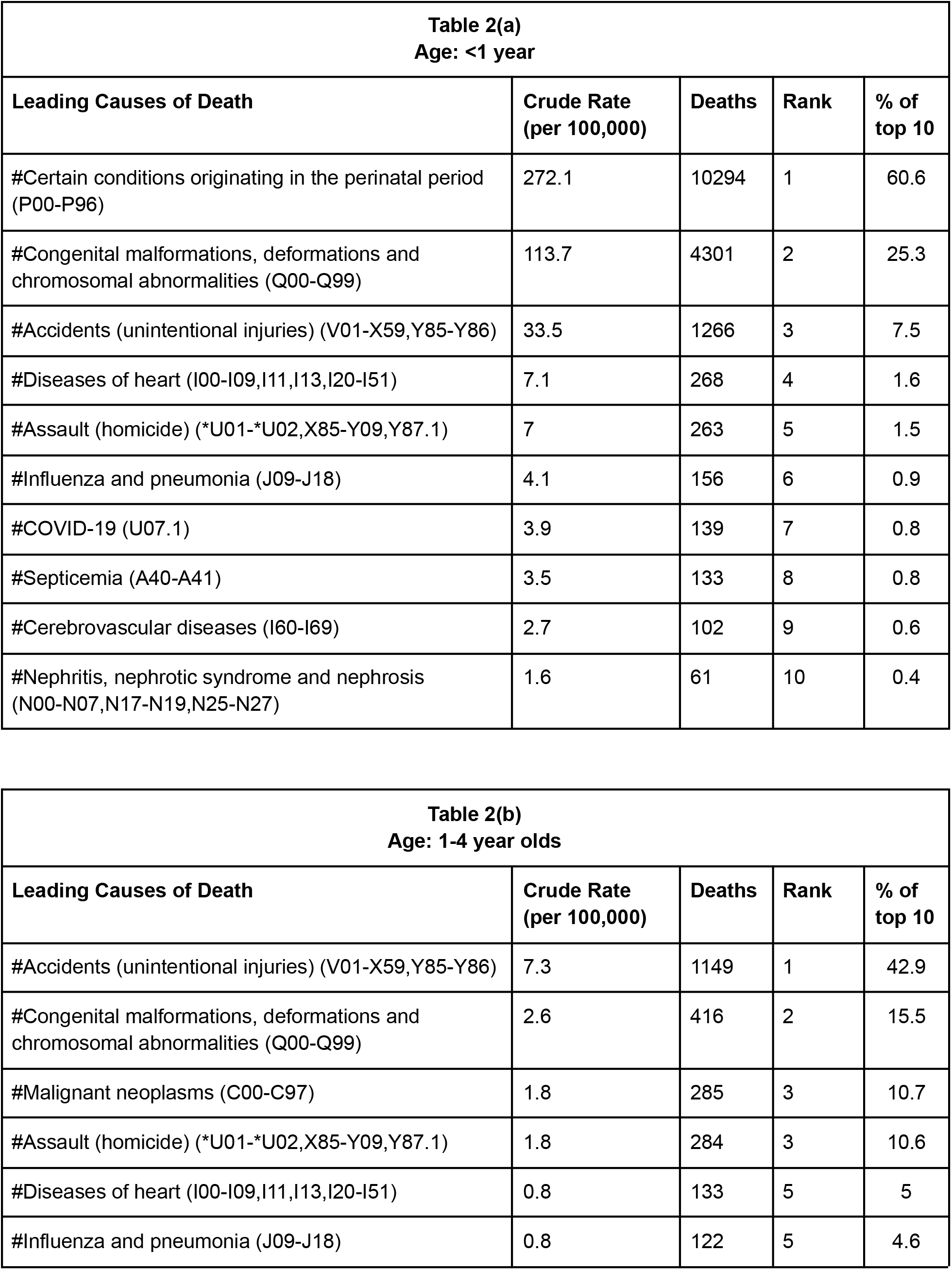
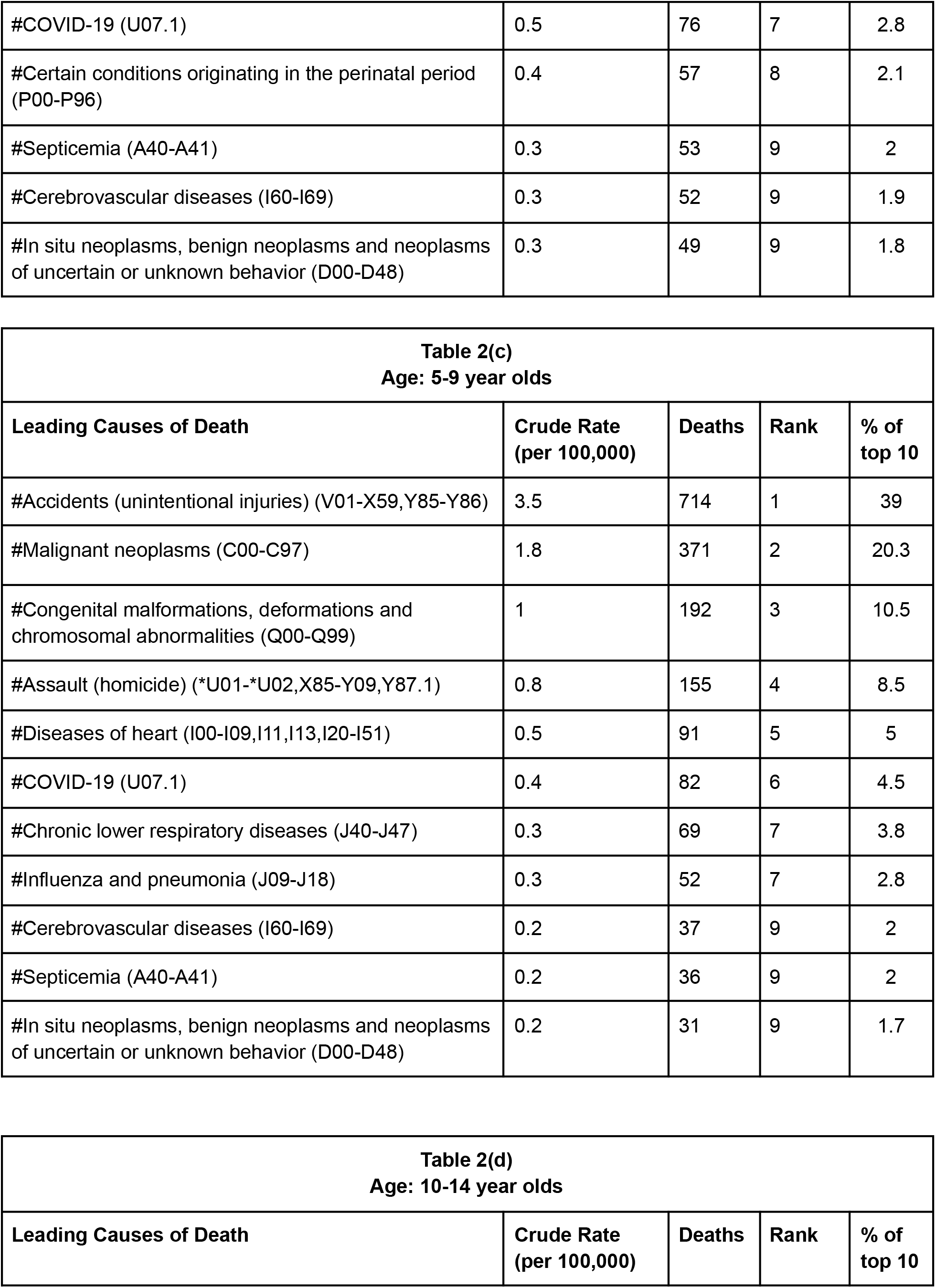
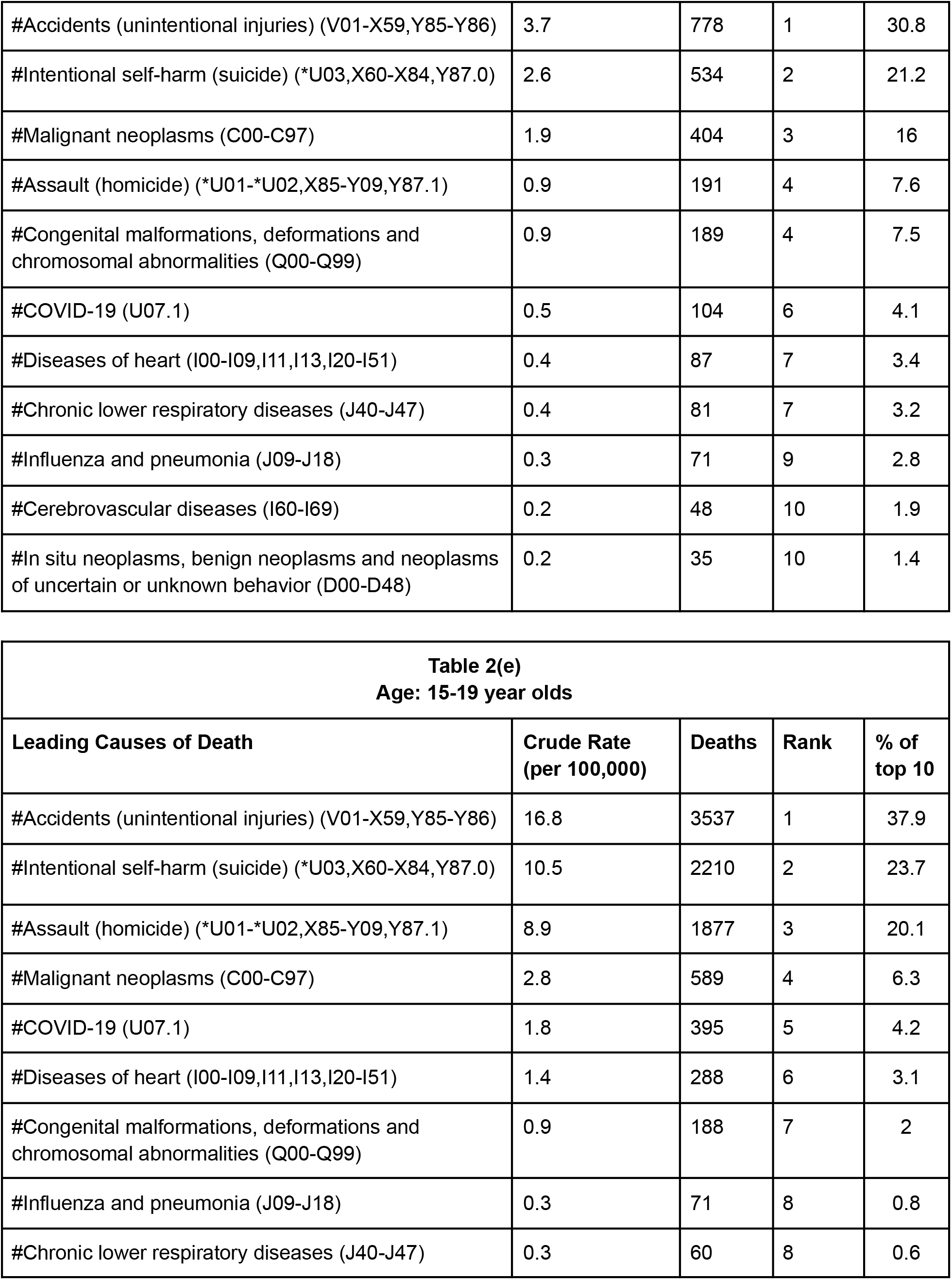
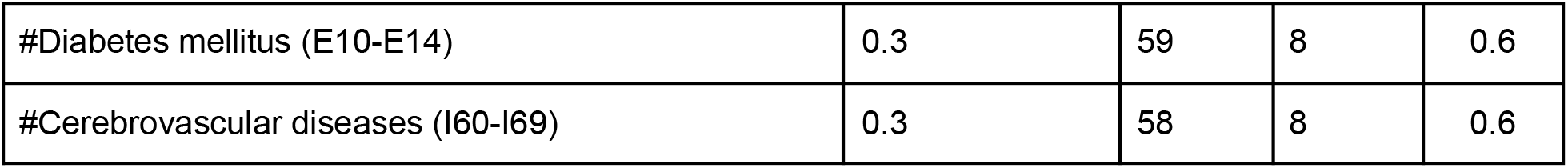
Leading causes of death from the rankable causes on the NCHS 113 Selected Causes of Death List, for children and young people aged 0-19 years in 2019 in the US ranked, compared to Covid-19 deaths (April 1, 2021-March 31, 2022). Deaths, crude rates per 100,000, ranks, and percentage of the 10 leading causes are shown for ages < 1 year (a), 1-4 years (b), 5-9 years (c), 10-14 years (d), 15-19 years (e). In all cases, Covid-19 is among the 10 leading causes of death. (Versions of Table 2(a) using the NCHS 130 Selected Causes of Infant Death are included in Supplementary Tables 1-3. The predominance of perinatal causes of death means that in the first 28 days of life, Covid-19 is not a leading cause of death, but it is a top 10 cause of death from ages 28-364 days.)

Excluding causes of death unrelated to disease (accidents, assault and suicide), Covid-19 ranked as the 5th leading cause of death in US CYP aged 0-19 years (Table 1). Considering infectious and respiratory diseases only, Covid-19 ranked as the top (1st) cause of death in US CYP aged 0-19 years (Table 3), followed by influenza and pneumonia as the 2nd leading cause (note that the causative agents of pneumonia may include multiple pathogens).

**Table 3.** Causes of death from the A00-B99 (Certain infectious and parasitic diseases) and J00-J98 (Diseases of the respiratory system) causes on the NCHS 113 Selected Causes of Death. Data is for children and young people aged 0-19 years in 2019 in the US ranked by number of deaths, compared to Covid-19 deaths (April 1, 2021-March 31, 2022). Deaths, crude rates per 100,000, and ranks are shown. Covid-19 is the top (1st) cause of death among infectious disease / diseases of the respiratory system. The NCHS 113 Selected Causes of Death can be grouped into rankable causes of death, indicated by the “#” symbol. Thus, categories overlap in the table, e.g. pneumonia (ICD-10 codes J12-J18) is a subset of the rankable cause #Influenza and pneumonia (J09-J18). Covid-19 was added as a rankable cause to the NCHS 113 Selected Causes of Death list in 2020^3^.

| **Table 3**<br>**Ages: 0-19 year olds** |  |  |  |
| --- | --- | --- | --- |
| **Leading Cause of Death from:**<br>**A00-B99 (Certain infectious and parasitic diseases) and**<br>**J00-J98 (Diseases of the respiratory system)** | **Crude Rate**<br>**(per 100,000)** | **Deaths** | **Rank** |
| #COVID-19 (U07.1) | 1.0 | 796 | 1 |
| #Influenza and pneumonia (J09-J18) | 0.6 | 472 | 2 |
| Other and unspecified infectious and parasitic diseases and their sequelae<br>(A00,A05,A20-A36,A42-A44,A48-A49,A54-A79,A81-A82,A85.0-A85.1,A85.8,A86-B04,B06-B09,B25-B49,B55-B99,U07.1) | 0.5 | 432 | 3 |
| Other diseases of respiratory system (J00-J06,J30-J39,J67,J70-J98) | 0.5 | 421 | 3 |
| Pneumonia (J12-J18) | 0.4 | 300 | 5 |
| #Septicemia (A40-A41) | 0.3 | 287 | 6 |
| #Chronic lower respiratory diseases (J40-J47) | 0.3 | 259 | 6 |
| Certain other intestinal infections (A04,A07-A09) | 0.3 | 223 | 6 |
| Asthma (J45-J46) | 0.2 | 206 | 9 |
| Influenza (J09-J11) | 0.2 | 172 | 9 |

Note that for consistency, we have used the NCHS 113 Selected Causes of Death (which was designed for ages 1+) for all CYP age group disaggregations and combinations, rather than the NCHS 130 Selected Causes of Infant Death. Perinatal causes of death predominate for neonates and thus Covid-19 is not a leading cause of death in the first 28 days of life using the 130 Causes List (Supplementary Table 1). Restricting to the age group 28 days - 364 days using the 130 Causes List, Covid-19 is a top 10 leading cause of death (Supplementary Table 2).

Our study period, April 1, 2021-March 31, 2022, coincides with substantial Delta and Omicron waves (Figure 1b). As a sensitivity analysis, we considered every 12 month period from April 2020 to July 2022 (Figure 1b). In the pre-Delta period, before July 2021, Covid-19 death rates were considerably lower than in the Delta and Omicron periods. Nevertheless, Covid-19 in this period would have ranked as the 9th leading cause of death in 2019, rather than the 8th leading cause of death.

## Conclusion

Early in the Covid-19 pandemic, comparisons of Covid-19 disease severity between age groups were a vital tool for appropriately allocating limited resources and prioritizing vaccination campaigns. However, long-term public health planning and management needs to be informed by the leading causes of deaths within each age group, a practice in the US dating back at least seven decades, beginning with the first publication of a leading cause of death ranking in 1952^2^. The most recent comprehensive data pre-pandemic on leading causes of death covers 2019^2^. Comparing to this period, we have shown that Covid-19 was a leading cause of death in CYP aged 0-19 years in the US for the period April 1, 2021-March 31, 2022, ranking 8th among all causes of deaths (comprising 2.4% of the 10 leading causes). Considering other 12 month periods during the pandemic does not qualitatively change our findings.

While other causes of death, such as accidents (22.5%), assault (8.4%) and suicide (8.3%) comprise a large percentage of the 10 leading causes of death, Covid-19 ranks 5th in disease-related causes of deaths (excluding accidents, assault and suicide), and 1st in deaths caused by infectious and respiratory diseases. Comparing deaths from Covid-19 with deaths from other vaccine-preventable diseases historically, Covid-19 caused substantially more deaths than major vaccine-preventable diseases did before vaccines became available: hepatitis A (3 deaths in children per year in the US), rotavirus (20-60 deaths), rubella (17 deaths), varicella (50 deaths)^11^.

Our findings need to be considered in the context of several limitations which mean that we may underestimate the true mortality burden of Covid-19 in CYP aged 0-19 years. Analyses of excess deaths have suggested under-reporting bias in Covid-19 deaths^12^; specific criteria for determining Covid-19 deaths is heterogeneous across the US and has changed over time; and delays to reporting may be substantial, especially for recent time periods. We consider Covid-19 as an underlying (and not contributing) cause of death only, but Covid-19 amplifies the severe impacts of other diseases and mortality hazards from co-infection (e.g. influenza^13^) are increased with accompanying comorbidities. The category of deaths from “influenza and pneumonia” combines a variety of causes, to which SARS-CoV-2 could be a contributing factor^13,14^. Recent evidence also suggests that Covid-19 may contribute to serious long-term sequelae^15^ in children and adolescents, which are unlikely to have been captured in these data.

In summary, we find that Covid-19 is now a leading cause of death for CYP aged 0-19 years in the US, and the top (1st) leading cause of death among infectious and respiratory diseases. Overall, deaths in CYP have increased over the Delta and Omicron waves compared to previous waves, likely reflecting the large numbers of CYP infected during these periods. A future variant or subvariant capable of displacing current Omicron subvariants will, by definition, have a growth advantage over Omicron, and there is no guarantee that its intrinsic severity will be reduced^16^. In the context of sustained transmission and circulation of SARS-CoV-2 in the US, the non-trivial risk posed by Covid-19 to CYP warrants utilization of a wide and robust array of tools to limit the extent of infection and severe disease in this age-group, through a combination of safe and efficacious vaccination against the disease^17^, appropriate pharmaceutical treatments, mitigations such as ventilation, and other non-pharmaceutical interventions.

## Data Availability

All data were originally obtained from https://wonder.cdc.gov/
and are available online at https://github.com/flaxter/covid19pediatric/

https://github.com/flaxter/covid19pediatric/

## Ethics

According to HRA decision tools (http://www.hra-decisiontools.org.uk/research/), our study is considered Research, and according to the NHS REC review tool (http://www.hra-decisiontools.org.uk/ethics/), we do not need NHS Research Ethics Committee (REC) review, as we only used (1) publicly available, (2) anonymized, and (3) aggregated data outside of clinical settings.

## Code Availability

All data and R scripts for replicating the results in this manuscript are available from: https://github.com/flaxter/covid19pediatric/

