## Supplementary Tables for "Covid-19 is a leading cause of death in children and young people ages 0-19 years in the United States"

| Ages: < 28 days | | | |
|---|---|---|---|
| **10 Leading Causes of Death (Infants)** | **Deaths** | **Crude Rate** | **Rank** |
| #Disorders related to short gestation and low birth weight, not elsewhere classified (P07) | 3374 | 0.9 | 1 |
| #Congenital malformations, deformations and chromosomal abnormalities (Q00-Q99) | 3047 | 0.8 | 2 |
| #Newborn affected by maternal complications of pregnancy (P01) | 1240 | 0.3 | 3 |
| #Newborn affected by complications of placenta, cord and membranes (P02) | 731 | 0.2 | 4 |
| #Bacterial sepsis of newborn (P36) | 579 | 0.2 | 4 |
| #Respiratory distress of newborn (P22) | 409 | 0.1 | 6 |
| #Neonatal hemorrhage (P50-P52,P54) | 335 | 0.1 | 6 |
| #Intrauterine hypoxia and birth asphyxia (P20-P21) | 320 | 0.1 | 6 |
| #Necrotizing enterocolitis of newborn (P77) | 312 | 0.1 | 6 |
| #Atelectasis (P28.0-P28.1) | 230 | 0.1 | 6 |

**Supplementary Table 1.** Leading causes of death from the rankable causes on the NCHS 130 Selected Causes of Infant Death List, for neonates (< 28 days old) in 2019 in the US ranked. There were 36 Covid-19 deaths (April 1, 2021-March 31, 2022) for this age group. Deaths, crude rates per 100,000 live births, and ranks are shown.

| Ages: 28-364 days | | | |
|---|---|---|---|
| **10 Leading Causes of Death (Infants)** | **Deaths** | **Crude Rate** | **Rank** |
| #Congenital malformations, deformations and chromosomal abnormalities (Q00-Q99) | 1254 | 0.3 | 1 |
| #Accidents (unintentional injuries) (V01-X59) | 1125 | 0.3 | 1 |
| #Sudden infant death syndrome (R95) | 1110 | 0.3 | 1 |
| #Diseases of the circulatory system (I00-I99) | 326 | 0.1 | 4 |
| #Assault (homicide) (*U01,X85-Y09) | 247 | 0.1 | 4 |
| #Diarrhea and gastroenteritis of infectious origin (A09) | 180 | 0 | 5 |
| #Influenza and pneumonia (J09-J18) | 156 | 0 | 5 |
| #Chronic respiratory disease originating in the perinatal period (P27) | 148 | 0 | 5 |
| #Septicemia (A40-A41) | 131 | 0 | 5 |
| #Disorders related to short gestation and low birth weight, not elsewhere classified (P07) | 71 | 0 | 5 |

**Supplementary Table 2.** Leading causes of death from the rankable causes on the NCHS 130 Selected Causes of Infant Death List, for infants 28-364 days old in 2019 in the US ranked. There were 103 Covid-19 deaths (April 1, 2021-March 31, 2022) for this age group. Deaths, crude rates per 100,000 live births, and ranks are shown.

| Ages: 0-364 days | | | |
|---|---|---|---|
| **10 Leading Causes of Death (Infants)** | **Deaths** | **Crude Rate** | **Rank** |
| #Congenital malformations, deformations and chromosomal abnormalities (Q00-Q99) | 4301 | 1.1 | 1 |
| #Disorders related to short gestation and low birth weight, not elsewhere classified (P07) | 3445 | 0.9 | 2 |
| #Accidents (unintentional injuries) (V01-X59) | 1266 | 0.3 | 3 |
| #Sudden infant death syndrome (R95) | 1248 | 0.3 | 3 |

| | | | |
|---|---|---|---|
| #Newborn affected by maternal complications of pregnancy (P01) | 1245 | 0.3 | 3 |
| #Newborn affected by complications of placenta, cord and membranes (P02) | 742 | 0.2 | 6 |
| #Bacterial sepsis of newborn (P36) | 603 | 0.2 | 6 |
| #Respiratory distress of newborn (P22) | 424 | 0.1 | 8 |
| #Diseases of the circulatory system (I00-I99) | 406 | 0.1 | 8 |
| #Necrotizing enterocolitis of newborn (P77) | 354 | 0.1 | 8 |

**Supplementary Table 3.** Leading causes of death from the rankable causes on the NCHS 130 Selected Causes of Infant Death List, for infants 0-364 days old in 2019 in the US ranked. There were 139 Covid-19 deaths (April 1, 2021-March 31, 2022) for this age group. Deaths, crude rates per 100,000 live births, and ranks are shown.
